# Insights into X-Linked Susceptibility to Parkinson’s Disease in the South African Population

**DOI:** 10.64898/2025.12.23.25342891

**Authors:** Kathryn Step, Emily Waldo, Thiago Peixoto Leal, Marla Mendes, Soraya Bardien, Ignacio F Mata, the Global Parkinson’s Genetics Program (GP2)

**Author notes:** **Correspondence to:** Professor Ignacio Mata, Genome Sciences and Systems Biology, Cleveland Clinic Research, Cleveland Clinic Foundation, Cleveland, OH, United States. **Target journal:** MDS.

## Abstract

X chromosome-wide association studies (XWAS) have successfully identified risk loci on the X chromosome associated with Parkinson’s disease (PD) susceptibility. However, only three such studies have been completed to date. Here, we present the first XWAS using an African cohort, comprising 690 PD cases and 826 controls. We applied an established XWAS workflow to perform male- and female-stratified analyses, as well as a combined meta-analysis. The male-stratified analysis identified five significant variants, including one lead locus (rs200539602), while the female-stratified analysis revealed 29 significant variants and two lead loci (rs2499550 and rs58045540), where rs2499550 is an upstream variant of the protein-coding gene *FAAH2*. The remaining female-stratified significant variants are expression quantitative trait loci for *SPIN2A, SPIN2B*, and *SPIN3*, which are highly expressed in the brain and nerve tissues, making them strong candidates for further investigation. One previously reported PD XWAS locus (rs28602900) was also replicated at a significance threshold of 0.05. The meta-analysis identified five variants surpassing chromosome-wide significance, including two lead loci (rs140715059 and rs141026964), the latter has no significant expression quantitative trait locus information but lies closest to the protein-coding gene *MAGEC2*, which may warrant further follow-up. None of the meta-analysis signals replicated in prior neurodegenerative disease XWAS. Overall, this study provides novel insights into the contribution of the X chromosome to PD susceptibility and represents the first PD XWAS to include participants of African ancestry, highlighting the importance of extending genetic studies to diverse populations.

## Introduction

Parkinson’s disease (PD) is a common movement and neurodegenerative disorder, with 11.7 million cases worldwide in 2021.^1^ PD is a complex disease influenced by a combination of genetic predispositions, lifestyle, and environmental exposures, with additional risk factors including age and sex.^2,3^ PD displays a strong sex bias, disproportionately affecting males, with an approximate male to female prevalence ratio of 1.18, however this varies across populations.^3^ Also, females are more frequently misdiagnosed, which may contribute to the underestimation of prevalence in women.^4–7^ Although genetic factors contribute substantially to PD risk, most efforts to identify susceptibility variants have relied on genome-wide association studies (GWAS). However, GWAS approaches target autosomal variants primarily and often exclude the sex chromosomes from analysis. Moreover, only approximately 0.05% of GWAS studies in the NHGRI-EBI GWAS Catalog have associations that include the X chromosome (Xchr).^8,9^ While there are various reasons for this exclusion,^10^ the Xchr carries over 800 protein-coding genes and accounts for approximately 5% of the human genome.^9^

There are several factors to consider when including the Xchr in genetic analysis, particularly when conducting association studies.^10^ These include the difference in inheritance of the Xchr between the sexes, with males only inheriting a maternal copy of the Xchr, while females inherit one Xchr from each parent.^11,12^ Moreover, in females, one of the two Xchr will undergo X-inactivation in each cell in a random manner to balance gene expression levels.^9,13^ However, this inactivation is not complete as some genes escape X-inactivation, leading to potential dosage differences that may influence a disease phenotype.^14^ In addition, there is no recombination during meiosis affecting the male Xchr. For this reason, the Xchr is more sensitive to evolutionary events that drive sequence variation.^11^

Recent research has highlighted the influence of sex in both the development of the brain as well as its physiology, showing the possibility of a sex bias in predisposition, severity, response to treatment, and disease progression, including in the context of neurodegenerative diseases.^5,15^ In the context of PD, the disparity of prevalence between sexes may be driven by either genetic sex or sex hormones impacting the biological pathways implicated in PD pathogenesis. Previous linkage analysis studies using multiplex families have identified a region on the Xchr (Xq21-q25) associated with PD.^16^

Despite this growing recognition of sex differences in PD, the contribution of sex chromosomes, particularly the Xchr, remains largely understudied. Only a handful of studies have investigated the link between the Xchr risk loci and neurological disorders, with only three PD X chromosome-wide association studies (XWAS) completed to date.^12,17,18^ Here, we aim to conduct the first PD XWAS using an African study collection to investigate the possible contribution of Xchr loci to disease aetiology.

## Methods and materials

An overview of the methods is provided in **Figure 1**. Briefly, the XWAS comprises four main steps (1) autosomal QC, (2) Xchr QC, (3) sex-stratified regression analysis, and (4) meta-analysis for combined results.

**Figure 1.**
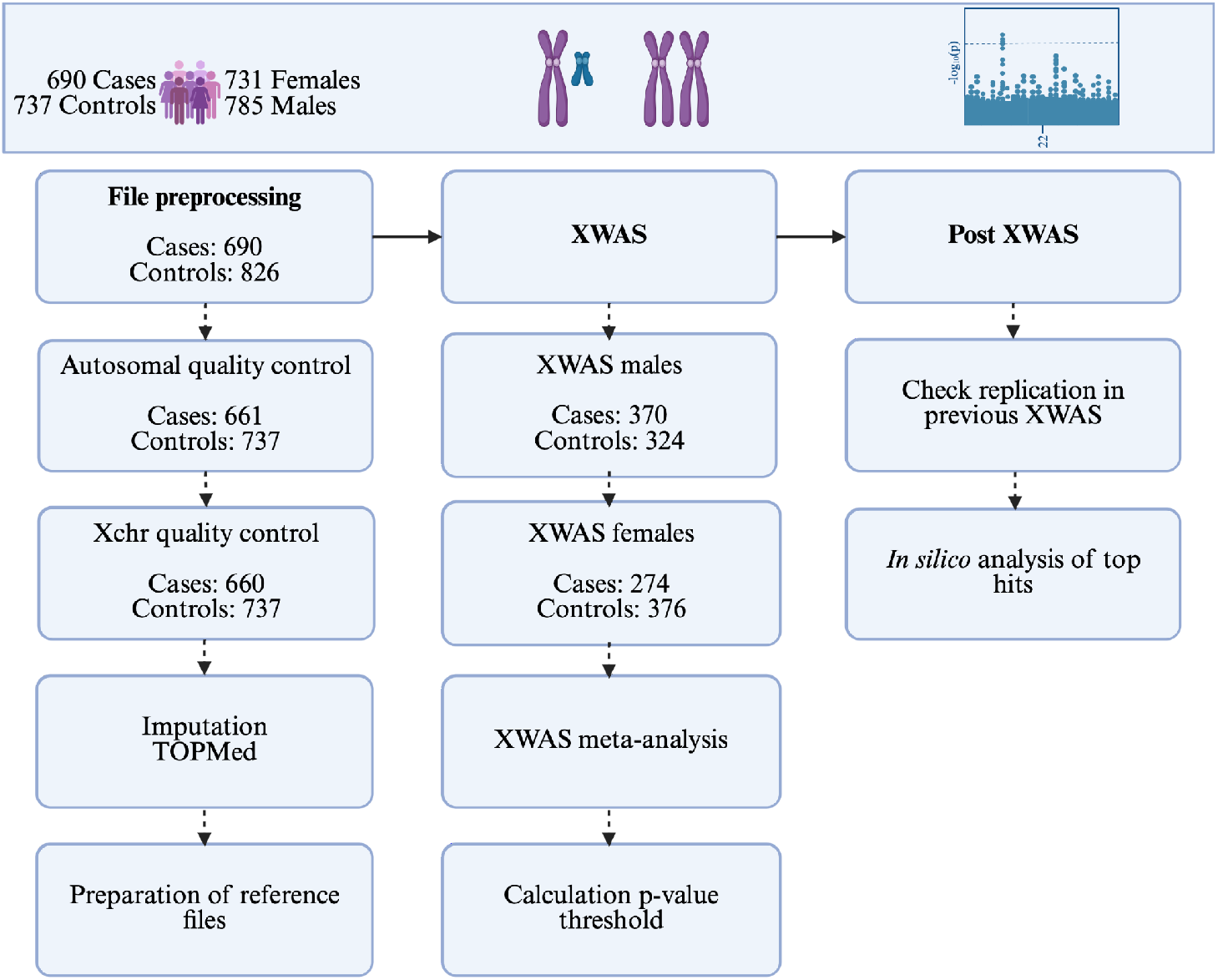
Overview of the layout of the study and the methods used. The number of cases and controls indicate how many individuals were retained at that step of the analysis. Xchr, X chromosome; XWAS, X chromosome-wide association study.

### Overview of the study cohort

The discovery cohort included 690 PD cases (293 females) and 826 controls (438 females). These participants were recruited as part of the South African Parkinson’s Disease Study Collection (Stellenbosch University Health Research Ethics Committee, 2002C/059). The PD cases were diagnosed according to the Queen’s Square Brain Bank Criteria,^19^ as previously described.^20^ The age at recruitment for the PD cases ranges from 18 to 102 years, and 18 to 92 years for controls. The genotyping was performed through the Global Parkinson’s Genetics Program^21,22^ using the NeuroBooster array (v1.0, Illumina, San Diego, CA).^23^

### Quality control, imputation, and population structure analysis

Quality control (QC) for autosomes was performed as previously reported.^24,25^ Briefly, at a sample level, QC removed samples with poor call rates, sex discrepancies, and excess heterozygosity. At the variant level, QC included removing duplicate variants, filtering monomorphic variants, excluding problematic probe variants, filtering for Hardy-Weinberg equilibrium, removing variants with low call rates, and applying minor allele threshold filtering. King v2.2.9 was used to infer the relationship coefficient with a threshold of 0.0884 to indicate second-degree relatives, where related individuals were excluded, making use of NAToRA and PLINK v2.0.^26–28^ Due to the unique nature of Xchr inheritance, the QC process for the Xchr differs from standard QC for autosomes.^8^ For the present study, we used the Xchr QC methods previously reported by Leal *et al*., 2023 and any individual removed during the autosomal QC process was excluded before Xchr QC. For the Xchr, variant-level QC included removing structural variants, removing duplicated and monomorphic variants, filtering potential probe variants, testing Hardy-Weinberg equilibrium in unrelated individuals, filtering for genotype and sample missingness, performing sex checks, correcting heterozygous calls in males, assessing differential missingness between cases and controls, testing Hardy-Weinberg equilibrium in females, removing monomorphic variants again, and assessing differential missingness and minor allele frequency between males and females. For sample-level QC of the Xchr, the same filters for the autosomes were applied. Detailed QC thresholds and procedures have been previously reported.^17^ In total, 1,397 participants (660 cases and 737 controls) were retained for downstream analysis (**Supplementary Table 1**). On the Xchr, QC was initially applied to 41,133 variants, of which 26,380 passed QC and were retained for imputation.

Imputation was performed using the Trans-Omics for Precision Medicine (TOPMed) Imputation Server,^29^ filtering out poorly imputed variants using an Rsq value of 0.3.^30^ We phased our Xchr data with Eagle using the 1000 Genomes Project Phase III reference (--allowRefAltSwap) before imputation with TOPMed.^31,32^ A projected principal component analysis was conducted to investigate the population structure of the study cohort, using imputed variants that overlapped with a genotyped reference panel (**Supplementary Figure 1**).^33^ This was done to identify and remove population outliers, which may introduce bias in the association analysis. Furthermore, we used previously reported global ancestry data from the South African Parkinson’s Disease Study Collection to access autosomal ancestral proportions.^25^ Global ancestry inference was performed using ADMIXTURE v1.3.0,^34^ and the results were compared between female and male participants to determine whether significant differences existed in ancestral components by sex.

### X chromosome-wide association analysis

Firth’s logistic regression was utilized for the sex-stratified XWAS using PLINK v2.0,^28^ as previously described.^17^ To account for population structure, stepwise regression, including 50 principal components (PCs), was used to identify the most relevant PCs for inclusion as covariates. PCs were generated using only Xchr genotyped variants, autosomal PCs were not used in this analysis. Stepwise regression was performed separately for females and males (**Supplementary Table 2**). For the female-only and male-only XWAS analyses, the selected PCs and age were included as covariates, while sex was excluded. Additionally, a meta-analysis was performed to combine the sex-stratified results using GWAMA v2.2.2.^35^ The heterogeneity between the two sexes was tested using this approach, as previously described.^36^ Post-XWAS *in silico* analyses were conducted using Functional Mapping and Annotation (FUMA) v1.3.8,^37^ Ensembl Variant Effect Predictor v114,^38^ Open Targets Genetics v22.10,^39,40^ and the Genotype-Tissue Expression (GTEx) Portal v10.^41^

### Calculating the X chromosome significance threshold

For a standard GWAS, the genome-wide significance threshold is indicated by p-value <5×10^-8^ and a suggestive significance threshold of p-value <1×10^-5^.^42^ However, when conducting an XWAS, only one chromosome is assessed; therefore, the number of independent tests (independent SNPs) performed is significantly lower.^36^ To ascertain the appropriate significance threshold for the Xchr, we calculated the number of effective tests and divided it by 0.05 (**Supplementary Methods**).^43^ This was calculated for the two independent XWAS results (male-XWAS and female-XWAS).^17^ The more stringent threshold, in this case the p-value calculated for the males (p-value: 6.04×10^−5^), was used as the chromosome-wide significance threshold for the meta-analysis combining the sex-stratified results.

### Replication using previously published studies

For replication, we looked at the significant hits identified through previous European-based XWAS for neurodegenerative disorders,^10^ including two Alzheimer’s disease studies^44,45^ and one PD study.^12^ Summary statistics from these studies were accessed in full, and the corresponding odds ratios and p-values were extracted for each variant. Replication was assessed across sex-stratified analyses as well as the combined meta-analysis, using a significance threshold of 0.05. Additionally, we evaluated whether the previously identified significant variants from these studies replicated in our dataset at a significance threshold of 0.05.

## Results

After QC and the removal of population outliers, 694 males (n= 370 cases) and 650 females (n= 274 cases) were retained for the analyses. The resulting summary statistics for the sex-stratified analysis were used in the meta-analysis. Autosomal global ancestry estimates were comparable between males and females, indicating minimal sex-related differences in ancestral proportions (**Supplementary Table 3**).

### X chromosome-wide association analysis reveals novel risk variants associated with Parkinson’s disease

For the male-XWAS, five variants reached chromosome-wide significance, with rs200539602 (OR [95% CI]: 3.42 [1.93-6.08]; p-value: 2.78×10^−5^) identified as the independent lead variant using *in silico* tools (**Table 1** and **Figure 2A**). All five SNPs are intergenic variants in linkage disequilibrium with each other (**Supplementary Table 4**). Moreover, no protein-coding genes have been functionally linked to any of the significant variants. For the female-XWAS, 29 variants reached chromosome-wide significance (p-value: 9.62×10^−5^; **Supplementary Table 4**). Two independent lead SNPs were identified using *in silico* approaches (**Table 1** and **Figure 2B**), rs2499550 (OR [95% CI]: 2.03 [1.45-2.84]; p-value: 3.28×10^−5^) and rs58045540 (OR [95% CI]: 0.21 [0.10-0.43]; p-value: 2.42×10^−5^). The nearest protein-coding gene to rs2499550 is *FAAH2*, which is expressed in the brain, amongst other tissues.^46^ Moreover, *FAAH2* has been linked to X-linked recessive neuropsychiatric disorders, including intellectual disabilities and Autism spectrum disorder.^47^ The second lead locus, rs58045540, has not been functionally linked to a protein-coding gene. Furthermore, several significant variants are intergenic or intronic variants in the protein-coding genes *SPIN2A, SPIN2B*, and *SPIN3*, all of which are highly expressed in brain and nerve tissues.^41^ When comparing significant variants from the sex-stratified analyses, none were shared between males and females.

**Figure 2.**
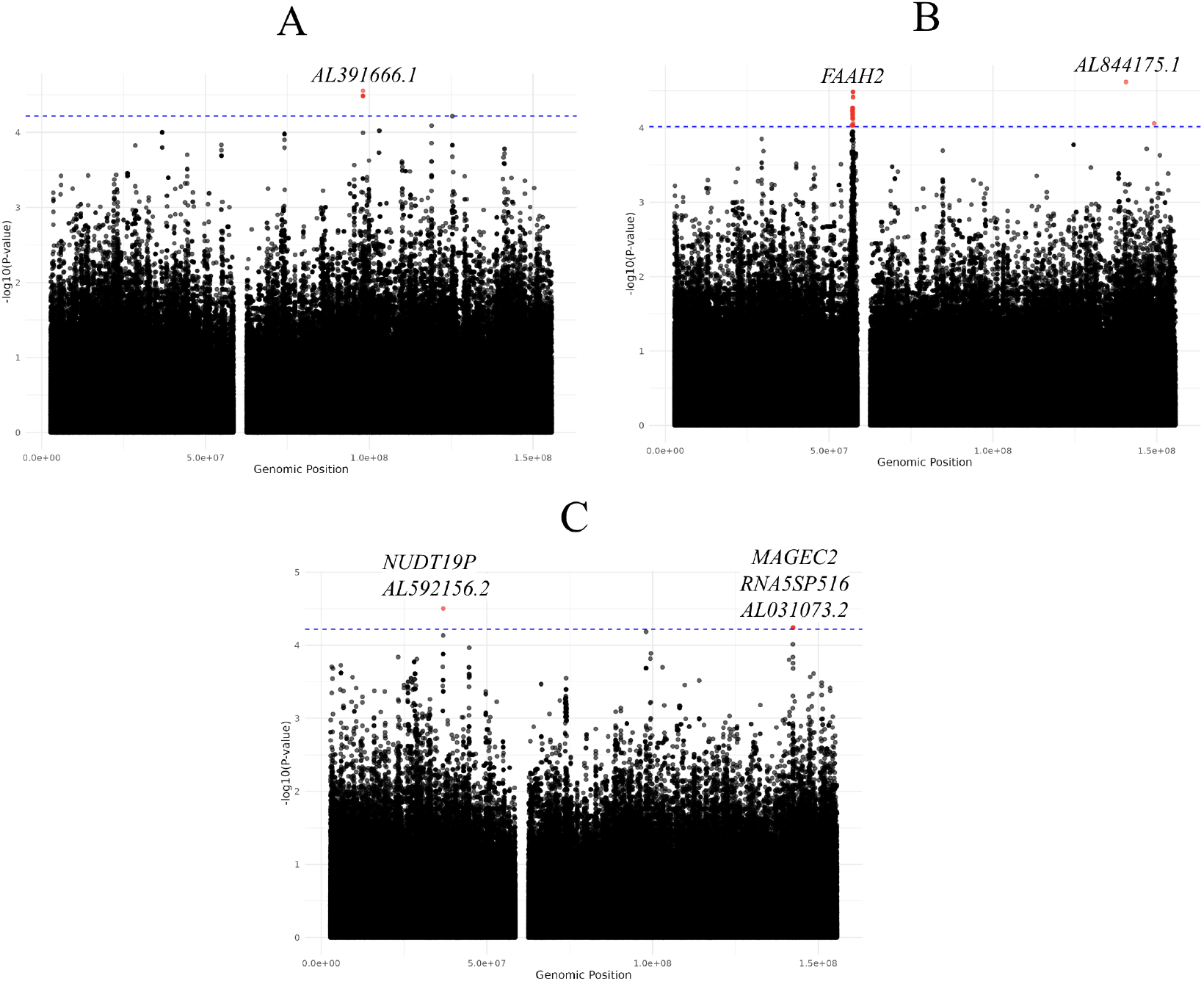
Manhattan plot of XWAS results using Xchr PCs. (A) For males with p-value significance p-value: 6.04×10^−5^, (B) For females with p-value: 9.62×10^−5^, and (C) Meta-analysis for males and females with p-value significance p-value: 6.04×10^−5^.

**Table 1.** Variants at chromosome-wide significance for the X chromosome-wide association analyses.

| Variant ID | P-value | Odds ratio (95% CI) | Nearest gene(s) |
| --- | --- | --- | --- |
| **Sex-stratified XWAS: Males** |  |  |  |
| rs200539602 | 2.78E-05 | 3.421 (1.925-6.081) | *AL391666.1* |
| **Sex-stratified XWAS: Females** |  |  |  |
| rs2499550 | 3.28E-05 | 2.033 (1.454-2.841) | *FAAH2* |
| rs58045540 | 2.42E-05 | 0.207 (0.100-0.430) | *AL844175.1* |
| **Meta-analysis** |  |  |  |
| rs141026964 | 5.73E-05 | 3.213 (1.820-5.672) | *MAGEC2*<br>*RNA5SP516*<br>*AL031073.2* |
| rs140715059 | 1.68E-05 | 2.973 (1.811-4.881) | *NUDT19P*<br>*AL592156.2* |
| Legend: CI, Confidence interval; XWAS, X chromosome-wide association study; Xchr, X chromosome |  |  |  |

### Meta-analysis of sex-stratified analysis identified Parkinson’s disease risk variants

For the meta-analysis, five variants reached chromosome-wide significance (p-value: 6.04×10^−5^; **Supplementary Table 4** and **Figure 2C**). Two independent lead loci were identified in this analysis (**Table 1**), rs140715059 (OR [95% CI]: 2.97 [1.81-4.88]; p-value: 1.68×10^−5^) and rs141026964 (OR [95% CI]: 3.21 [1.82-5.67]; p-value: 5.73×10^−5^). The significant SNPs are intronic, non-coding, or downstream of a gene. The nearest protein-coding gene to rs141026964 is *MAGEC2*, whereas rs140715059 is not located near any annotated protein-coding gene. Furthermore, there was no overlap between the significant variants identified in the meta-analysis and those in the sex-stratified analyses (**Supplementary Table 4**).

### Replication of multiple loci in female-stratified analysis

Three previously published neurodegenerative XWAS were included in the replication analysis for this study.^12,44,45^ However, none of the significant variants replicated in either of the previous Alzheimer’s disease XWAS. Replication was observed only with the first PD XWAS conducted by Le Guen and colleagues.^12^ No replication signals were observed in the male-stratified XWAS or meta-analysis across studies. However, replication was detected in the female-stratified analysis, where 23 variants replicated in the previously published PD XWAS (**Table 2**).^12^ Notably, one of the two female lead loci, rs2499550, was replicated (OR: 0.963; p-value: 0.004). The two lead loci reported in the XWAS by Le Guen and colleagues are not in linkage disequilibrium with the variants identified as significant in the present study. However, rs28602900 (OR: 1.558; p-value: 0.018) did replicate in the female-stratified dataset at the 0.05 threshold. This locus was previously replicated in a PD XWAS by Leal and colleagues in 2023.^17^

**Table 2.**
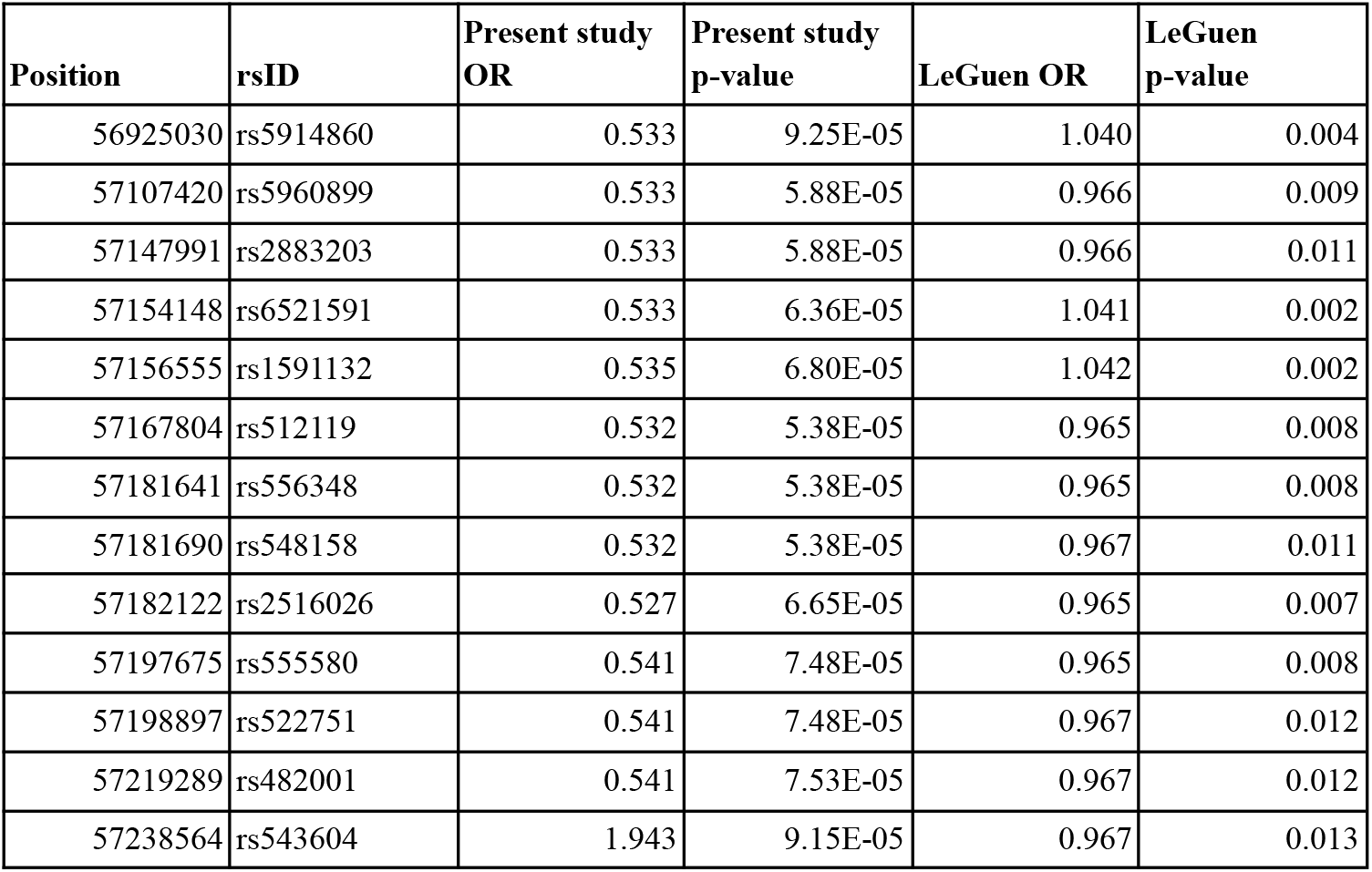
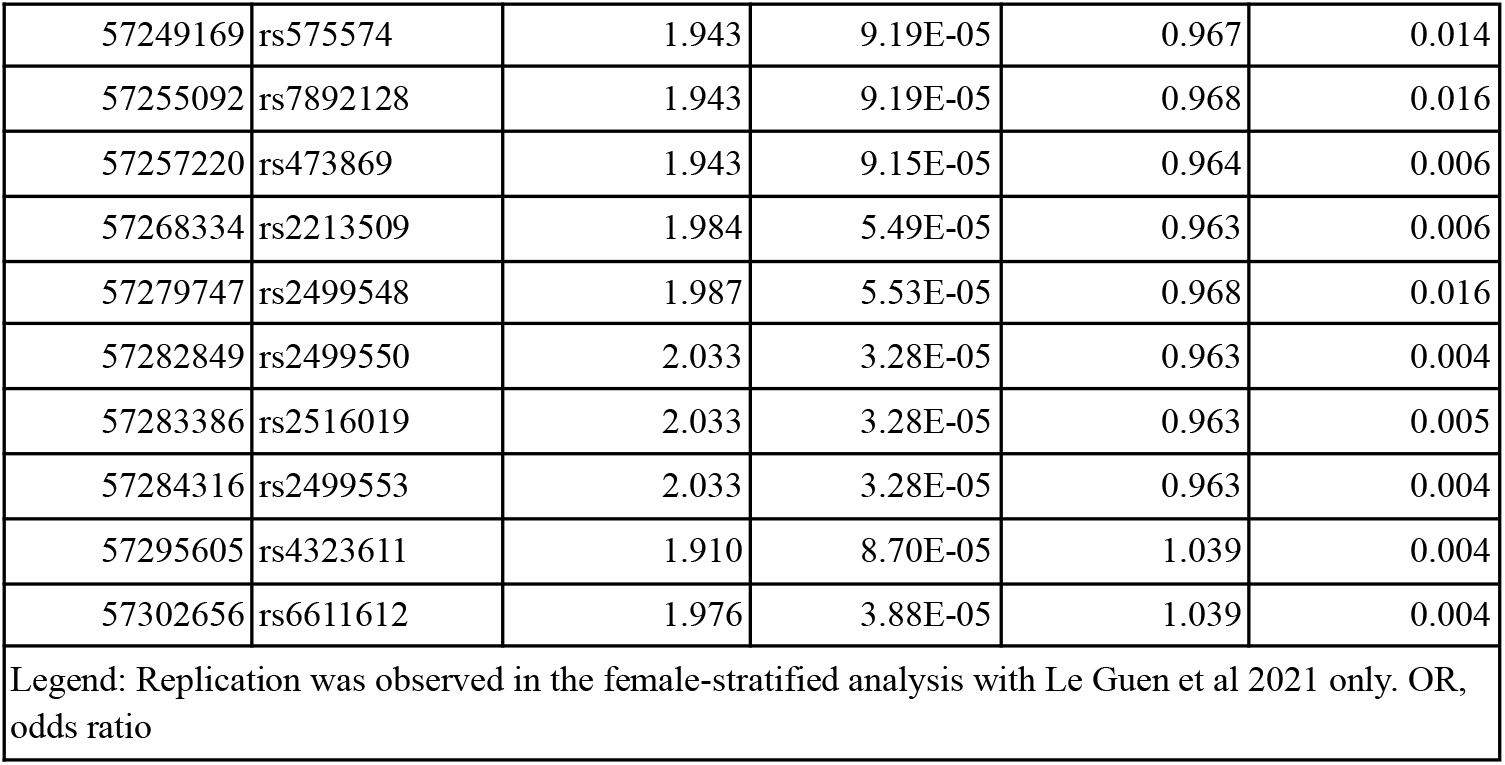
Variants replicated at nominal p <0.05 from previous X chromosome-wide association studies.

## Discussion

To our knowledge, this study represents the first XWAS for PD that included individuals of African ancestry and uniquely incorporates participants from the South African population. Although the Xchr spans approximately 156 million base pairs, it is routinely excluded from genome association studies due to its unique biology and the additional analytical challenges it presents.^9,10^ Here, we utilized an established and improved XWAS workflow by Leal and collaborators and applied it to a uniquely admixed African population.^17^

The female-stratified analysis identified 29 significant variants with two lead loci (rs2499550 and rs58045540). Of particular interest is rs2499550, which the aforementioned *in silico* analyses classify as a variant of uncertain significance and map as an expression quantitative trait locus (eQTL) upstream of the protein-coding gene *FAAH2*. This gene is expressed in the brain and has been implicated in X-linked recessive neuropsychiatric disorders.^47^ Functional studies in patient-derived fibroblasts have shown that variants in *FAAH2* can reduce enzymatic activity, while affected individuals exhibit neurological features including motor impairment, gait abnormalities, and tremor.^47^ Inhibition of the FAAH enzyme has been shown in preclinical models to improve parkinsonian motor symptoms,^48^ highlighting the potential functional relevance of the *FAAH2* variants identified in this study. Furthermore, the other significant variants are eQTLs for *SPIN2A, SPIN2B*, and *SPIN3*, all of which are highly expressed in the brain and nerve tissues, as determined by the GTEx Analysis Release V10 (dbGaP Accession phs000424.v10.p2) on 11/20/25, making these loci strong candidates for further investigation in the context of PD. Notably, one previously reported lead locus from Le Guen *et al*. (rs28602900) replicated in our dataset at a significance threshold of 0.05.^12^ This XWAS locus has been associated with a significant brain putamen volume reduction in the UK Biobank dataset.^12^ Moreover, this variant was replicated in a previous PD XWAS on a Latin American cohort,^17^ highlighting its association with PD across populations.

The meta-analysis, which combined the sex-stratified results, revealed five variants surpassing chromosome-wide significance and two lead loci, rs140715059 and rs141026964. The latter has no significant eQTL information, but lies closest to the protein-coding gene *MAGEC2*, which may warrant further investigation. This gene is involved in ubiquitin ligase activity, a pathway implicated in PD pathogenesis.^49,50^ The male-stratified analysis identified five significant variants, including one lead locus (rs200539602). None of the variants in the male-stratified or meta-analysis replicated in previous neurodegenerative disease XWAS, indicating that they currently lack supporting evidence from earlier studies. Although these findings highlight potential novel risk factors for PD, *in silico* functional assessment provided limited evidence for their biological relevance, underscoring the need for further experimental validation to clarify their role in disease pathophysiology.

This study contributes to our understanding of Xchr susceptibility factors for PD, particularly within the South African population. To date, three previous PD XWAS have been published. In 2021, Le Guen *et al* published the first PD XWAS, including 296,685 individuals (16,521 cases) of European ancestry.^12^ Their study identified two loci of interest (rs7066890 and rs28602900),^10,12^ where the latter was replicated in our female-stratified analysis presented above. In 2023, Leal *et al* published the first PD XWAS to include individuals of admixed ancestry, focusing on a Latin American cohort of 1,481 individuals (798 cases).^17^ This study identified eight Xchr regions associated with PD and successfully replicated the rs28602900 variant from the 2021 PD XWAS.^10,17^ In 2025, Liao *et al* published the first XWAS for PD progression, analyzing 4,467 PD cases of European ancestry.^18^ They reported 11 independent variants associated with cognitive decline in PD cases.^18^ Overall, these findings highlight Xchr susceptibility variants and sex differences in the genetic architecture of PD, aligning with the findings presented in the present study.

Although this study has several strengths and has identified novel risk loci, there are several limitations. These include the relatively small sample size, particularly in the sex-stratified analysis, which resulted in several peaks falling just below chromosome-wide significance and may have prevented detection of true associations that a larger cohort could reveal. Also, the absence of a South African reference panel contributed to the identification of 36 (n= 20 female) individuals as population outliers during PCA, which further reduced statistical power. While a more appropriate reference panel would not eliminate ancestry outliers entirely, it would likely reduce their number by providing a better fit to the underlying population structure. Additionally, there are no ancestry-matched XWAS summary statistics available for replication. Another limitation is the restricted ability and accuracy of *in silico* tools, especially for variants on the Xchr, to investigate XWAS hits and ascertain their biological relevance. Finally, this study did not include functional validation of the identified variants, which limits our understanding of their biological relevance and potential role in PD.

In conclusion, this study applies recently updated XWAS methodologies to provide novel insights into the contribution of the Xchr to PD. We further validated our findings by comparing them with previously published XWAS of neurodegenerative disorders, and notably, we replicated one of the previously reported PD XWAS loci (rs28602900) in our dataset. Importantly, this work represents the first PD XWAS to include individuals of African ancestry, addressing a critical gap in genetic research. While further studies are needed to fully elucidate Xchr susceptibility variants in PD, our findings advance the characterization of genetic risk and help broaden the current understanding of PD genetics across diverse populations.

## Data Availability

Data used in the preparation of this article were obtained from Global Parkinson's Genetics Program (GP2). GP2 is funded by the Aligning Science Across Parkinson's (ASAP) initiative and implemented by The Michael J. Fox Foundation for Parkinson's Research (https://gp2.org). For a complete list of GP2 members see https://doi.org/10.5281/zenodo.7904831. The full summary statistics are available with Tier 1 access through the Accelerating Medicines Partnership in Parkinson's Disease (AMP-PD) (https://www.amp-pd.org/). The summary statistics for the replication cohorts can be obtained as follows: NHGRI-EBI GWAS catalog (https://www.ebi.ac.uk/gwas/) for Le Guen et al., 2021 (GCST90104085) and Le Borgne et al., 2024 (GCST90449045), and Simmonds et al., 2024 is available at https://github.com/UKDRI/XWAS_AD_summary_stats. The quality control pipeline is available at https://github.com/MataLabCCF/XWAS. The association analysis pipeline is available at https://github.com/MataLabCCF/XWAS_v2 and the NAToRA pipeline is available at https://github.com/ldgh/NAToRA_Public. The pipelines were developed and are maintained by Dr. Thiago Peixoto Leal and are available at https://github.com/MataLabCCF. Additionally, an overview of the analysis and any additional scripts not available through the Mata Lab GitHub can be found in the GP2 public domain on GitHub (https://github.com/GP2code/SouthAfrican_XWAS; DOI 10.5281/zenodo.18021965).

https://www.ebi.ac.uk/gwas/

https://github.com/UKDRI/XWAS_AD_summary_stats

## Author contributions

K.S., S.B., and I.M. conceptualized the manuscript. K.S. wrote the first manuscript draft. T.P.L. developed the workflows for the data analysis. M.M.d.A developed scripts for the data analysis. E.W. assisted with the data analysis. All authors reviewed, edited, and approved the final version of the manuscript for submission.

## Declaration of interests

I.F.M. has received honorarium from the Parkinson’s Foundation PD GENEration Steering Committee and Aligning Science Across Parkinson’s Global Parkinson Genetic Program (ASAP-GP2).

## Acknowledgements

We would like to acknowledge and thank the study participants for their contribution. The data used for the analysis was obtained from the GP2 (https://gp2.org) which is funded by the Aligning Science Across Parkinson’s (ASAP) initiative and implemented by The Michael J. Fox Foundation for Parkinson’s Research (https://michaeljfox.org/). A complete list of GP2 members is available at https://gp2.org. All figures were created using BioRender (https://www.biorender.com/). We thank Kate Andersh, Laurel Screven, and Natalia López González del Rey for their work as scientific project managers for this project. We also acknowledge the Centre for High Performance Computing (CHPC), South Africa, for providing computational resources. All figures were created using BioRender.com. *For open access, the author has applied a CC BY public copyright license to all Author Accepted Manuscripts arising from this submission*.

## Funding

K.S. is supported by The Michael J. Fox Foundation and Aligning Sciences Across Parkinson’s Disease Global Parkinson Genetic Program. I.M. is supported by the National Institutes of Health (1R01NS112499, U01AG076482, R01NS132437), The Michael J. Fox Foundation and the Aligning Science Across Parkinson’s Global Parkinson Genetic Program (ASAP-GP2), American Parkinson’s Disease Association (APDA) and Department of Veterans Affairs (I01BX005978-01A1). He also receives honorarium for his participation in Parkinson’s Foundation PD GENEration Steering Committee and Aligning Science Across Parkinson’s Global Parkinson Genetic Program (ASAP-GP2) Operations Committee. The Michael J. Fox Foundation (MJFF-026283 for E.W. and I.F.M.) and Alzheimer’s Disease Sequencing Project (ADSP) (5U01AG076482-03 for E.W.). T.P.L is supported by National Institutes of Health (1R01NS112499) and Department of Veterans Affairs (I01BX005978-01A1). S.B. received support from the National Research Foundation of South Africa (grant number 129429),the South African Medical Research Council (Self-Initiated Research Grant) and the Centre for Tuberculosis Research (CTR) of the South African Medical Research Council.

## Notes

### Author Declarations

Data used in the preparation of this article were obtained from Global Parkinson's Genetics Program (GP2). GP2 is funded by the Aligning Science Across Parkinson's (ASAP) initiative and implemented by The Michael J. Fox Foundation for Parkinson's Research (https://gp2.org). For a complete list of GP2 members see https://doi.org/10.5281/zenodo.7904831. The full summary statistics are available with Tier 1 access through the Accelerating Medicines Partnership in Parkinson's Disease (AMP-PD) (https://www.amp-pd.org/). The summary statistics for the replication cohorts can be obtained as follows: NHGRI-EBI GWAS catalog (https://www.ebi.ac.uk/gwas/) for Le Guen et al., 2021 (GCST90104085) and Le Borgne et al., 2024 (GCST90449045), and Simmonds et al., 2024 is available at https://github.com/UKDRI/XWAS_AD_summary_stats. The quality control pipeline is available at https://github.com/MataLabCCF/XWAS. The association analysis pipeline is available at https://github.com/MataLabCCF/XWAS_v2 and the NAToRA pipeline is available at https://github.com/ldgh/NAToRA_Public. The pipelines were developed and are maintained by Dr. Thiago Peixoto Leal and are available at https://github.com/MataLabCCF. Additionally, an overview of the analysis and any additional scripts not available through the Mata Lab GitHub can be found in the GP2 public domain on GitHub (https://github.com/GP2code/SouthAfrican_XWAS; DOI 10.5281/zenodo.18021965). The individual-level data has be de-identified.

